# HIV BG505 SOSIP.664 trimer with 3M-052-AF/alum induces human autologous tier-2 neutralizing antibodies

**DOI:** 10.1101/2024.05.08.24306957

**Authors:** William O Hahn, K. Rachael Parks, Mingchao Shen, Gabriel Ozorowski, Holly Janes, Lamar Ballweber-Fleming, Amanda S Woodward Davis, Chris Duplessis, Mark Tomai, Antu K Dey, Zachary K. Sagawa, Stephen C De Rosa, Aaron Seese, Latha Kallur Siddaramaiah, Leonidas Stamatatos, Wen-Hsin Lee, Leigh M Sewall, Dalton Karlinsey, Hannah L Turner, Vanessa Rubin, Sarah Furth, Kellie MacPhee, Michael Duff, Lawrence Corey, Michael C Keefer, Srilatha Edupuganti, Ian Frank, Janine Maenza, Lindsey R Baden, Ollivier Hyrien, Rogier W. Sanders, John P. Moore, Andrew B Ward, Georgia D Tomaras, David C Montefiori, Nadine Rouphael, M Juliana McElrath

**Affiliations:** Vaccine and Infectious Disease Division, Fred Hutchinson Cancer Center, Seattle, Washington; Department of Integrative Structural and Computational Biology, The Scripps Research Institute, La Jolla, California; Division of AIDS, National Institute of Allergy and Infectious Diseases, Bethesda, Maryland; 3M Healthcare, St. Paul, Minnesota; IAVI, New York, New York; Access to Advanced Health Institute, Seattle, Washington; Department of Laboratory Medicine and Pathology, University of Washington, Seattle, Washington; University of Rochester, Department of Medicine, Rochester, New York; Department of Medicine, Division of Infectious Diseases, Emory University School of Medicine, Atlanta, Georgia; School of Medicine, University of Pennsylvania, Philadelphia; Department of Medicine, University of Washington, Seattle, Washington; Brigham and Women’s Hospital, Boston, Massachusetts; Academic Universities Medical Center, Amsterdam, the Netherlands; Weill Cornell Medicine, New York, New York; Center for Human Systems Immunology and Departments of Surgery and Integrative Immunobiology, Duke University School of Medicine, Durham, North Carolina; Department of Surgery, Duke University, North Carolina; Hope Clinic, Emory University, Atlanta, Georgia

## Abstract

Stabilized trimers preserving the native-like HIV envelope structure may be key components of a preventive HIV vaccine regimen to induce broadly neutralizing antibodies (bnAbs). We evaluated trimeric BG505 SOSIP.664 gp140, formulated with a novel TLR7/8 signaling adjuvant, 3M-052-AF/Alum, for safety, adjuvant dose-finding and immunogenicity in a first-in-healthy adult (n=17), randomized, placebo-controlled trial (HVTN 137A). The vaccine regimen appeared safe. Robust, trimer-specific antibody, B-cell and CD4+ T-cell responses emerged post-vaccination. Five vaccinees developed serum autologous tier-2 nAbs (ID50 titer, 1:28-1:8647) after 2-3 doses targeting C3/V5 and/or V1/V2/V3 Env regions by electron microscopy and mutated pseudovirus-based neutralization analyses. Trimer-specific, B-cell-derived monoclonal antibody activities confirmed these results and showed weak heterologous neutralization in the strongest responder. Our findings demonstrate the clinical utility of the 3M-052-AF/alum adjuvant and support further improvements of trimer-based Env immunogens to focus responses on multiple broad nAb epitopes.

**KEY TAKEAWAY/TAKE-HOME MESSAGES:** HIV BG505 SOSIP.664 trimer with novel 3M-052-AF/alum adjuvant in humans appears safe and induces serum neutralizing antibodies to matched clade A, tier 2 virus, that map to diverse Env epitopes with relatively high titers. The novel adjuvant may be an important mediator of vaccine response.

## Introduction

Development of an effective preventive vaccine is essential to achieve long-term immunity against HIV-1 acquisition. A successful HIV vaccine will likely need to induce potent broadly neutralizing antibodies (bnAbs) to contemporary circulating viral strains, which is supported by findings from nonhuman primate studies and human efficacy trials administering HIV-1 bnAbs as monoclonal antibodies (Gautam et al., 2018; Pauthner et al., 2019; Saunders et al., 2022; Corey et al., 2021). No candidate HIV vaccine regimen evaluated has induced detectable serum bnAbs, and thus efforts to design and evaluate HIV-1 envelope (Env) immunogens that can elicit these responses is a major priority.

Most previously tested HIV immunogens express HIV-1 Env as monomeric proteins or viral vector inserts, which do not mimic the native-like envelope (Env) spike that mediates HIV-1 entry into target cells (Guttman et al., 2015; Kwon et al., 2015; Pancera et al., 2014; Kwong et al., 2020). As a result, the elicited antibodies are limited in their ability to neutralize any viral strains beyond lab-adapted tier 1 viruses and cannot neutralize tier 2 or tier 3 viruses, which are reflective of circulating viral strains (Guttman et al., 2015; Kwon et al., 2015; Pancera et al., 2014; Kwong et al., 2020; Montefiori et al., 2018). Thus, neutralizing epitopes conserved among contemporary tier 2 and 3 strains or their epitope precursors are not exposed on these immunogens as seen in natural infection.

To address this challenge, recombinant stabilized HIV-1 envelope glycoprotein trimers have been engineered through extensive efforts to modify the Env trimer sequence in order to maintain a more natural conformation (Klasse et al., 2013; Khayat et al., 2013; Sanders and Moore, 2017; Ward and Wilson, 2017; Dey et al., 2018). The BG505 SOSIP gp140, an early prototype stabilized trimer, has aided in the identification of bnAb structural interactions with target Env epitopes and more recently has provided a platform for HIV-1 designs. Preclinical studies have demonstrated that BG505 SOSIP.664 gp140 can induce neutralizing antibodies in rabbits, nonhuman primates, and cows, which can recognize its tier-2 autologous HIV-1 clade A strain, BG505, and in some cases protect against challenge with the matched strain (Schooten et al., 2021; Sanders et al., 2015; Pauthner et al., 2017; de Taeye et al., 2015; Havenar-Daughton et al., 2016) Here we report an early clinical trial in humans, HVTN 137 Part A, to analyze the safety, specificities and neutralization activities of antibodies induced with the clinical grade BG505 SOSIP.664 trimer.

While structural modifications help maintain a native-like Env trimeric structure of BG505 SOSIP.664, substantial antibody responses directed against the gp41 base of soluble trimers have been detected following immunization in both preclinical studies and clinical trials (Houser et al., 2022; Kulp et al., 2017; Hu et al., 2015). The base epitopes lack the glycan shield that surrounds other regions of the trimer and are not exposed on intact HIV-1 during natural infection, and antibodies recognizing the base epitopes are incapable of neutralizing HIV (Turner et al., 2021). The extent to which base-binding antibody responses preclude other, more physiologically relevant antibodies in humans is an important question for the HIV vaccine field because of the high immunologic barriers to generating antibodies capable of neutralizing diverse HIV strains. In preclinical vaccine models, B cells recognizing base epitopes predominate and may restrict formation of more desirable B cell lineages from the germinal center (Abbott and Crotty, 2020). The first clinical study evaluating a BG505 SOSIP.664 trimer with an additional disulfide bond and formulated with Alum adjuvant reported only the induction of base-directed serum antibodies with no evidence of tier 2 virus neutralization (Houser et al., 2022). Moreover, many target bnAb precursor B cells exist at low frequencies and are likely to be subdominant during vaccination. Thus, a more potent adjuvant formulated with the trimer could potentially support the expansion of sub-dominant B cell clones that are ultimately necessary for neutralization (Kasturi et al., 2020; Oleszycka and Lavelle, 2014; Ciabattini et al., 2016; Knudsen et al., 2016).

In HVTN 137A, we selected the 3M-052 TLR7/8 agonist, representing a novel class of promising vaccine adjuvants, to formulate with the BG505 SOSIP.664. The synthetic imidazoquinolinone 3M-052 is structurally related to R848 with modifications intended to minimize systemic distribution (Smirnov et al., 2011). Non-human primate vaccine studies using 3M-052 within nanoparticles or adsorbed to alum as an adjuvant with either BG505 SOSIP.664 gp140 trimer or HIV-1 1086.C gp140 Env monomer elicited high neutralizing antibody titers and increased antigen-specific lymph node germinal center activities (Kasturi et al., 2020). Moreover, modulating the strength of the GC interaction by adoptive transfer of CD4+ T cells may overcome known issues with immunodominance of Env trimers (Lee et al., 2020). With these insights, we designed the HVTN137A trial to determine whether BG504 SOSIP.664 in combination with 3M-052-AF + Alum is safe in humans and is capable of inducing antibodies that neutralize more wild-type like Env (tier 2) viruses and potent CD4+ T cells.

## Materials and Methods

### Study design and population

HVTN 137 Part A (Clinicaltrials.gov NCT04177355) is a multisite, randomized, controlled phase 1 clinical trial examining the safety and immunogenicity of BG505 SOSIP.664 gp140 formulated with 3M-052-AF (1 or 5 ug) adsorbed to aluminum hydroxide (500 µg) when administered intramuscularly. The HIV Vaccine Trials Network (HVTN) conducted and the National Institute of Allergy and Infectious Disease (NIAID), Division of AIDS (DAIDS) sponsored the trial. Fred Hutchinson Cancer Center Institutional Review Board (IRB, FHIRB0010314) approved the study protocol and procedures. All participants demonstrated understanding of the protocol and provided written informed consent prior to enrollment. Study participants were adult men and women not living with HIV-1 between the ages of 18 and 50 (**Table S1**). The protocol document (Clinicaltrials.gov NCT04177355) provides a full description of the inclusion and exclusion criteria and the step-wise enrollment for safety by group. To protect participant privacy, all participant identifiers reported in this manuscript are publication IDs generated by the research group which are not known to anyone outside the group.

### Vaccine products

cGMP-manufactured BG505 SOSIP.664 gp140 and the Tris-NaCl diluent, used for diluting the vaccine antigen and as placebo, was provided by IAVI (New York, NY) for clinical use. Aluminum hydroxide, referred to here as Alum, was provided by the NIAID Vaccine Research Center and sourced originally from Brenntag Biosector (Denmark) as Alhydrogel. 3M-052 was purchased from 3M Corporation and formulated into 3M-052 aqueous formulation (AF) by IDRI (now Access to Advanced Health Institute, Seattle, WA) (Fox et al., 2016).

### Enrollment, randomization and blinding

Group 1 participants were randomized 5:1 to receive either BG505 SOSIP.664 gp140 with 1µg of 3M-052-AF plus 500 µg Alum (n=5) or placebo (n=1). After a pre-planned hold and safety evaluation, Group 2 participants were enrolled and randomized to receive BG505 SOSIP.664 gp140 with either the 5µg dose of 3M-052-AF plus 500 µg Alum (n=10) or placebo (n=1).

### Clinical procedures

Participants received BG505 SOSIP.664 gp140, diluted to 100 µg dose with Tris-NaCl, and administered as a 1mL volume admixed with the adjuvant formulation into the deltoid muscle of the upper arm. Immunizations were scheduled at enrollment (month 0) and two months later, but due to COVID-19 pandemic disruptions, the actual vaccination schedule varied substantially. However, institutional-mandated, COVID-19-related schedule delays permitted review of preliminary results, which prompted a protocol amendment to offer an optional third dose to participants, planned to be given at month six (**Table S2**).

Participants were observed for at least 30 minutes following immunization, and they recorded in a diary any pre-defined solicited adverse events (reactogenicity) for seven days post-immunization. Unsolicited adverse events were recorded throughout participation in the trial. Adverse events were coded according to the Medical Dictionary for Regulatory Activities and graded for severity according to the DAIDS Table for Grading of the Severity of Adult and Pediatric Adverse Events, version 2.1. Participants were followed for twelve months after the final vaccination with in-person clinic visits. Serum and peripheral blood mononuclear cell (PBMC) samples isolated from blood samples were processed and cryopreserved <8 hours from venipuncture at protocol-specified timepoints.

## Laboratory Procedures

### Antibody neutralization

Serum neutralizing antibodies against HIV-1 were measured as a function of reductions in Tat-regulated luciferase (Luc) reporter gene expression in TZM-bl cells after a single round of infection with Env-pseudotyped viruses (Sarzotti-Kelsoe et al., 2014; Montefiori, 2009). Neutralization titer was defined as the serum inhibitory dilution (ID50/ID80) or monoclonal antibody inhibitory concentration (IC50/IC80) at which relative luminescence units (RLU) were reduced by either 50% or 80% relative to control wells containing only virus and cells, after subtraction of background RLU (cells only). Serum samples testing positive against BG505/T332N were assayed for neutralization against a global panel of 9 heterologous tier-2 viruses: 246-F3_C10_2, 25710-2.43, BJOX002000.03.2, CH119.10, Ce1176_A3, Ce703010217_B6, CNE55, TRO.11, and X1632-S2-B10 (deCamp et al., 2014; Mkhize et al., 2023). Pseudovirus mutants using BG505 were used as previously described (Klasse et al., 2018).

### Binding antibody multiplex assay (BAMA)

IgG specific serum antibodies against HIV-1 gp41 and BG505 SOSIP AviB antigens were measured on a Bio-Plex instrument (Bio-Rad) using a custom HIV-1 Luminex assay (Tomaras et al., 2008; Haynes et al., 2012; Yates et al., 2014). Standard positive controls consisting of predefined monoclonal antibodies with known specificity (PGT145, PGT141), and polyclonal sera from people living with HIV. Standard negative controls of serum from people living without HIV and the CH65 mAb were included at a single dilution. Serum samples were tested across six dilution points at a five-fold serial dilution, with 1:50 dilution as the starting point. The results are presented as an area under the mean fluorescence intensity (MFI) curve (AUC) across the 6 dilutions, calculated using the trapezoidal rule and with MFI values truncated at zero. A positive response met 3 criteria around increase in MFI from the baseline response, and absolute MFI at the 1:50 dilution.

### Antigen-specific T cell intracellular cytokine staining (ICS) and cluster analysis

A 27-color flow cytometry panel was used to identify HIV-1 specific CD4+ and CD8+ T cell responses using a validated ICS assay; details of the antibody staining reagents and protocol have been previously published (Horton et al., 2007; Cohen et al., 2021). Peptides (BioSynthesis, Inc.) derived from the BG505 SOSIP.664 gp140 sequence were synthesized as 15-mers overlapping by 11 amino acids and combined into BG505 gp120 (128 peptides) and BG505 gp41 (37 peptides) peptide pools. Thawed, then rested PBMC were stimulated for 6h with synthetic peptide pools. Unstimulated cells (including DMSO, the peptide diluent) were used as a negative control and cells stimulated with Staphylococcal enterotoxin B were used as a positive control. Samples were stimulated with peptide pools for BG505 gp41 and BG505 gp120, to determine HIV-specific responses and the CMV pp65 peptide pool as an antigen-specific control. The background (negative control) subtracted percentages of cells expressing IFN-γ and/or IL-2 in response to gp120 and gp41 peptide pools were summed for the total vaccine-induced response. Positivity determination was based on a participant-level statistical comparison of number of cytokine-producing cells for stimulated and unstimulated wells (Horton et al., 2007).

Analysis of the high-dimensional ICS dataset assays included: (1) clustering identification using the Leiden algorithm (the *leiden* R package with *n_iterations = 10*) (Traag et al., 2019); and (2) determining the frequency of antigen-stimulated cells in comparison to the negative control-stimulated cells using *MIMOSA* (Finak et al., 2014) for identification of significant T cell antigen-specific responses. A multiplicity adjustment across the number of peptide pools is made to the individual peptide pool p-values.

Data for all samples were used (all treatment groups) under the four stimulation conditions (BG505 gp41, BG505 gp120, and CMV peptide pools, and also the negative control) and over three time points two weeks post-immunization (M0.5, M2.5 and M6.5). One participant’s samples were removed because this individual has a CD45RA genetic variant that results in all cells being positive for that marker and thus the naïve and memory subsets cannot be identified. Cluster analysis was restricted to CD4+ T cells, gated as negative for other lineage markers (CD14, CD19, CD16, CD56, CD8), positive for CD3 and negative for the viability dye that labels dead cells (UVID). CD4+ T cells expressing at least 1 of 7 of the functional markers (IFN-γ, IL-2, TNF-α, CD154, IL-4, IL-5/13, and IL-17a) were extracted and used as input. Several data transformation methods were tested and we chose the arcsinh transformation (co-factor 500) with an asymmetrical transformation (root 3) to de-emphasize cells with highly negative (or positive) fluorescence intensity in the arcsinh transformed distribution.

Using this strategy, 18 clusters were identified. As noted above, MIMOSA positivity was performed for each cluster for the gp120, gp41 and CMV peptide pool stimulations to determine which clusters included responses above the negative control stimulation. Eight of the clusters had no positive responses, indicating that they are likely not specific to the peptide pools. One additional cluster contained many positive responses among placebo recipients with the marker profile suggesting potential staining artifact (low magnitude expression of IL-5 and/or IL-13 without other functional markers). Therefore, these 12 clusters were removed from further analysis. Data for the remaining 6 clusters were graphed as background-subtracted percentages of CD4+ T cells for the total Env response (gp120 and gp41 peptide pools summed).

### Antigen-specific B cell identification

Participant PBMCs were incubated with fluorescently-labeled recombinant BG505-SOSIP.664 Env proteins, kindly provided by JP Moore (Cornell-Weill Medicine), and other fluorescently labeled antibodies (Cohen et al., 2021). Live total B cells were defined based on viability dye staining, scatter profile and doublet exclusion. The following lineage markers were used to identify B cells: negative for CD3, CD56, and CD14, and positive for both CD19 and CD20. IgG+ B cells were further gated as IgD- and IgG+. The percent of BG505 SOSIP.664 gp140-specific IgG+ B cells, out of the total number of IgG+ B cells, was reported. A positive response determination was based on a participant-level statistical comparison of Env-specific B cells after vaccination vs. baseline using a Fisher’s exact test.

### B cell receptor sequence analysis

BG505 SOSIP.664 gp140-specific IgD-were sorted by flow cytometry. VH/VL genes were sequenced using the Chromium Next GEM Single Cell 5’ Kit (v2). 10x Genomics libraries were sequenced at the Fred Hutch genomics core, which provides FASTQ files demultiplexed by index. Matched gene expression, V(D)J and feature FASTQ files were then processed through the high computing cluster using 10x Genomic’s latest stable version of CellRanger (7.1.0) and the *multi* processing option. The resulting files were loaded into R using our proSCessoR package that leverages other R packages such as dsb^1^ and Seurat^2^ to clean up background signal and further demultiplex samples by cell hashtag, allowing identification of singlet events versus multiplet droplets. V(D)J heavy- and light-chain pairing utilized metrics such as read depth and UMI counts to determine the appropriate contig selection of available contigs for a given cell for final pairing. IMGT HighV-Quest was used for gene annotation and productivity determination, and productive paired cells proceeded to clonotype analysis using the Immcantation Change-O pipeline (Mulè et al., 2022; Stoeckius et al., 2018; Manso et al., 2021; Lefranc et al., 2015; Lefranc, 2014; Gupta et al., 2015). Analysis of annotated sequences was done in R using the tidyverse (Wickham et al., 2019) suite of tools.

### Monoclonal antibody Biolayer interferometry (BLI)

Monoclonal antibodies (IgG1 subclass) were produced by GenScript from paired VH/VL sequences recovered from BG505 SOSIP.664 gp140-specific B cells. All monoclonal antibodies were derived from VH/VL genes recovered from Participant 11 (**Table S5**). Sequences were selected for production as a monoclonal antibody based on being a member of a large clonal lineage and high levels of somatic hypermutation. Antibodies were diluted to 20µg/ml and captured on the Anti-hIgG Fc Capture Biosensors. Antibodies were evaluated for binding to the analyte, BG505 SOSIP.664 gp140 at 1uM.

### Electron Microscopy-based Polyclonal Epitope Mapping (EMPEM)

EMPEM was performed to identify Env epitope regions recognized by vaccine-induced serum antibodies and synthesized monoclonal antibodies. Polyclonal Fab preparation from serum and sample preparation for electron microscopy were previously described (Bianchi et al., 2018; Tas et al., 2022). Briefly, 1 mL serum from each study participant collected after two and three vaccine doses was heat inactivated. Total IgG was isolated using CaptureSelect IgG-Fc Multispecies Affinity Matrix (Thermo Fisher) and Fab digestion was performed using papain (Sigma Aldrich). 15 µg of BG505 SOSIP.664 gp140 were incubated with 1 mg of Fab mixture (which also contains Fc and residual papain) overnight and purified the next day using a Superose 6 Increase (Cytiva) gel filtration column. If later EM imaging suggested a high degree of antibody-induced trimer dissociation (Turner et al., 2021), complex formation was repeated using BG505-CC2IP.664 gp140, which contains an interprotomer disulfide (501C-663C) in place of the original 601C-605C intraprotomer disulfide. Purified complexes were diluted to 0.03 mg/mL and deposited on glow-discharged carbon coated copper mesh grids, followed by staining with 2% (w/v) uranyl formate. Imaging was performed on one of three instruments: a) FEI Tecnai Spirit T12 equipped with an FEI Eagle 4k x 4k CCD camera (120 keV, 2.06 Å/pixel), b) FEI Talos equipped with an FEI Ceta 4K CMOS camera (200 keV, 1.98 Å/pixel), or c) an FEI TF20 equipped with a TVIPS TemCam F416 CMOS 4k x 4k camera (120 keV, 1.67 Å/pixel). All data were processed using Relion 3.0 (Zivanov et al., 2018) using standard 2D and 3D classification procedures. Polyclonal Fab densities were compared to previously published high resolution cryoEM models (Antanasijevic et al., 2021) for epitope assignment. Composite maps were generated using UCSF Chimera (Zivanov et al., 2018). Representative maps have been deposited to the Electron Microscopy Data Bank as summarized in **Fig. S1**. Participants were numbered 1-17 which corresponds to PubIDs in **Table S2.**

### Statistical Analysis

Participants were analyzed according to randomization group, except that placebo recipients in Groups 1 and 2 were combined for analysis. Safety outcomes were evaluated among all enrolled participants. Immunogenicity outcomes after a given dose were evaluated among participants who received all doses up to that time point.

## Data availability

Most data underlying the figures and tables can be found in the paper or the online supplemental material. FCS files from flow cytometry studies are available from the corresponding author upon reasonable request. The EMPEM representative maps (**Fig. S1)** are available in Electron Microscopy Data Bank (https://www.emdataresource.org). The remainder of the data underlying this study are not publicly available due to potential privacy issues.

## Supplemental figures

Figure S1 displays EMPEM data of serum antibodies from all participants after the second and third vaccination. Figure S2A demonstrates serum neutralizing activities against tier 1 HIV-1 MW965.26. Figure S2B provides ID50 pseudovirus neutralizing antibody activities in serum from Participant 17 after the third dose, including responses to the autologous wildtype BG505 HIV and the panel of mutated pseudoviruses used to map measure heterologous tier 2 activity. Figure S3A displays Biolayer Inferometry binding strength of the indicated monoclonal antibodies to BG505 SOSIP.664. Figure S3B demonstrates the neutralization reduction in participant 17 observed with various mutant knockouts. Table S1 provides the study participant demographics. Table S2 demonstrates the study visit and vaccination schedule. Table S3 details the local reactogenicity that participants experienced following immunization. Table S4 describes the systemic reactogenicity that participants experienced following immunications. Table S5 describes the binding and neutralizing activities of each participant following immunization. Table S6 describes clonotype size and sequence features of B cell-derived monoclonal antibodies.

## Results and Discussion

### Enrollment and Safety Assessments

Since 3M-052-AF/Alum was administered for the first time as a vaccine adjuvant in humans, a primary objective of HVTN 137 Part A was to establish the safety of 3M-052-AF/Alum with a target dose of 5µg of the 3M-052-AF component. Alum was added to the 3M-052-AF formulation to collocate antigen and adjuvant (Fox et al., 2016). The vaccine regimen was deemed safe among the 17 participants with no unsolicited severe adverse events (SAEs); no related SAEs, no adverse events of special interest (including potentially immune mediated medical conditions), no deaths; and no unplanned study pauses. The original plan was to administer vaccine (n=15) or placebo (n=2) on the day of enrollment (month 0) and the second dose on day 56 (month two). However, disruptions associated with the COVID-19 pandemic led to an adjusted, heterogenous vaccine administration schedule with an optional third dose (**Tables S2**).

Although the vaccine was generally tolerable, a small number of participants did experience local reactogenicity, including two grade 3 local reactogenicity events in the 5µg dose group (**FIg. 1, Table S3**). The onset of these events was around day 7. The course was similar to delayed local induration and redness observed following mRNA vaccines for COVID-19 (Baden et al., 2021; Chapin-Bardales et al., 2021). At no point was the pain/tenderness greater than mild, and this did not impact activities of daily living. One participant discontinued further administration and the other participant received a second dose uneventfully. Most participants reported only mild to moderate symptoms (**Table S4**). There were slightly more systemic reactogenicity events in the 5µg group as compared to the 1µg dose group (**Fig. 1)**. The overall rate of Grade 3 systemic reactogenicity (6 episodes per 35 doses) is similar to currently licensed vaccines that are and considered to be reactogenic, including an 18% rate of grade 3 systemic reactogenicity following the second dose of Spikevax, the COVID-19 mRNA vaccine (Moderna) (Baden et al., 2021) and 9% rate for the Shingrix vaccine with AS01_b_ adjuvant (Cunningham, 2016; Baden et al., 2021). Other than the delayed, persistent erythema in one recipient in the 5µg dose recipient described above, all reactogenicity symptoms resolved within 14 days, with the majority generally resolving within 7 days. Based on these findings, we conclude that the vaccine is safe but reactogenic.

**Figure 1.**
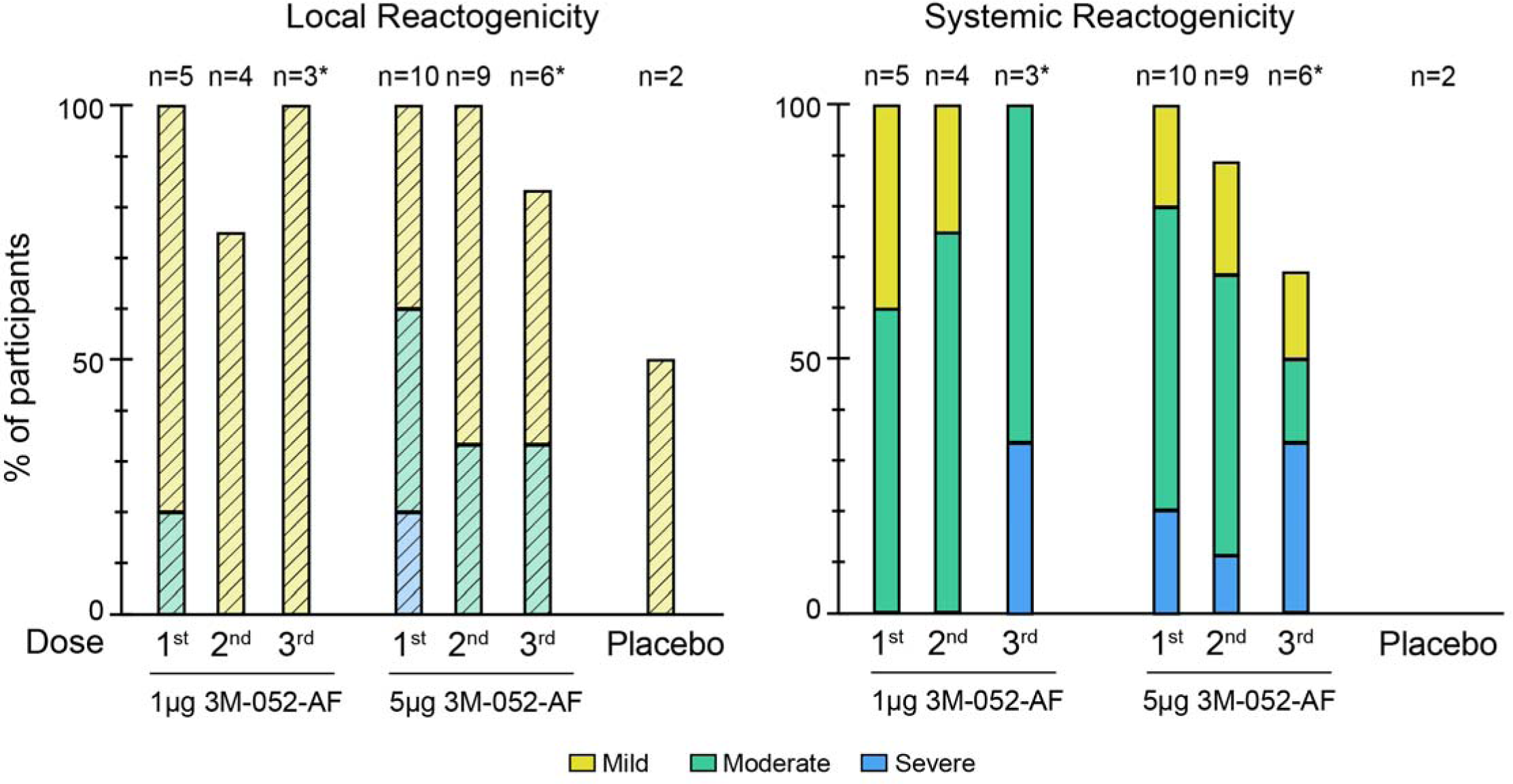
Local and systemic reactogenicity associated with BG505 SOSIP.664 with 3M-052-AF + Alum. The maximum local (left) and systemic (right) events reported within seven days after the 1^st^, 2^nd^, and optional 3^rd^ dose (indicated by *) of BG505 SOSIP.664 gp140 with 3M-052-AF (at 1µg or 5µg) + Alum or in the Placebo group. Reactions were graded as mild (yellow), moderate (green) or severe (blue). Number of participants in each group are listed above.

### BG505 SOSIP.664 with 3M-052-AF + Alum elicits strong antibody responses, including autologous tier 2 neutralizing activities

Overall, the vaccine formulation elicited robust HIV Env binding and neutralizing antibody responses (**Fig. 2A and B, Table S5**). Higher IgG binding antibody response rates were observed after the second immunization in the 5mcg group (9/10 participants) than in the 1mcg group (2/5 participants). However, all participants mounted IgG antibody responses to BG505 SOSIP.664 following the third immunization (**Fig. 2A**). Median binding antibody titers (AUC) after the second immunization (month 2.5) were 1591.6 (Group 1) and 6673.9 (Group 2), and after the third immunization (month 6.5) were 17214.3 (Group 1, n=3) and 18029.1 (Group 2, n=5). Of note, only one participant (137-118) in the 5 mcg dose group had detectable IgG antibodies binding to BG505 SOSIP.664 gp41 (ID50 AUC 1243) after the second immunization.

**Figure 2.**
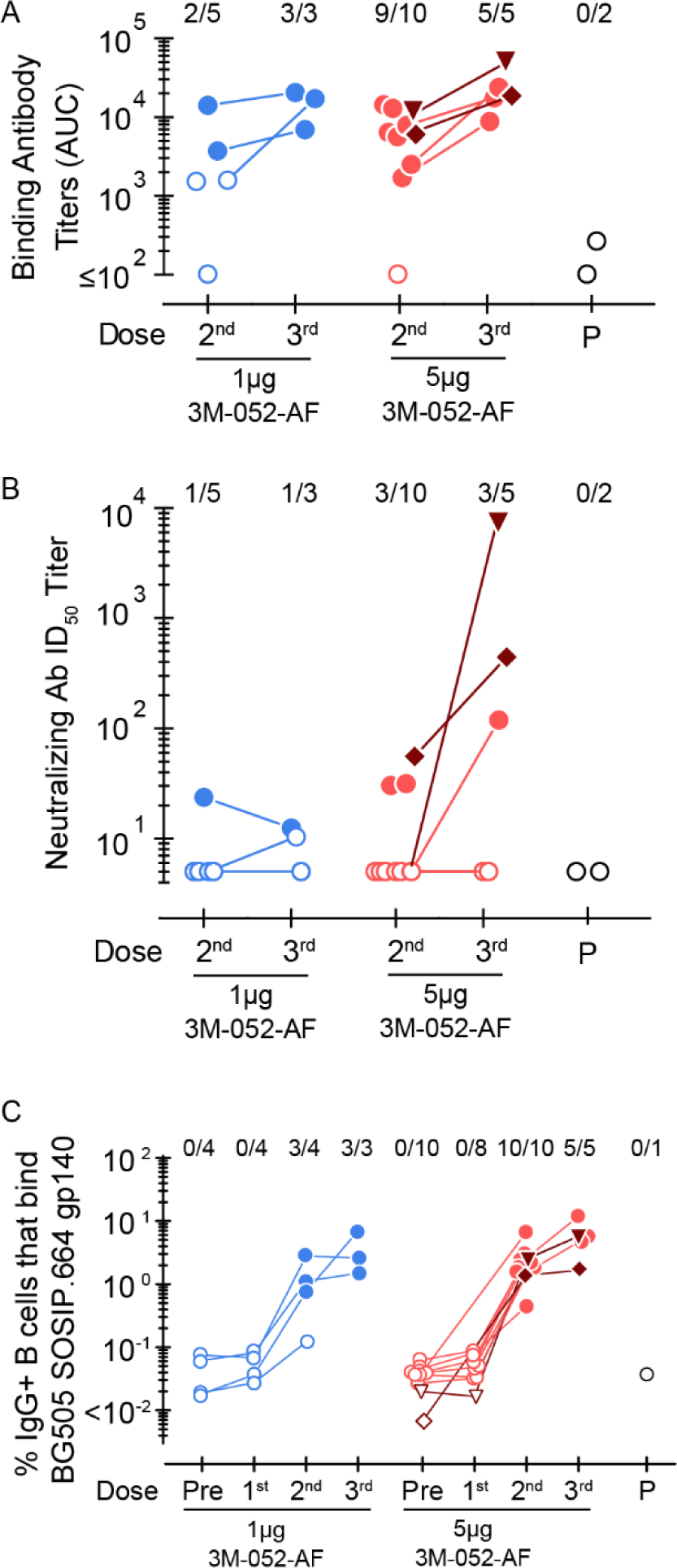
Vaccine-induced serum binding antibodies, tier 2 neutralizing antibodies and circulating memory IgG+ B cells. (A) BG505 SOSIP.664 gp140 specific responses were measured by binding antibody multiplex assay (BAMA) in serum two weeks following the indicated dose of vaccine. IgG binding magnitude area under the curve (AUC) values are shown against BG505 SOSIP.664 gp140. B) ID_50_ neutralizing antibody titers against the autologous tier 2 HIV-1 BG505/T332N using the TZMbl neutralization assay after the indicated vaccine dose. (C) Percent of IgG+ B cells that bind BG505 SOSIP.664 gp140 fluorescent probe as assessed by flow cytometry. B cells were identified using doublet exclusion, lymphocyte scatter profile, viability dye and stained negative for lineage markers CD3, CD56 and CD14; positive for CD19 and CD20. IgG+ B cells are gated as IgD- and IgG+. For the x axis, “Pre” is the Month 0 sample taken before vaccination, and “P” represents the placebo group. The dark red lines indicate responses of the same two individuals with the highest neutralizing titers after the third dose, shown in B (participant 11 indicated by ▼ and Participant 17 indicated by ♦). Filled symbols indicate positive responses.

To examine anti-viral function, serum antibody neutralizing activities were assessed to a reference HIV tier 1 strain (MW965.26) (**Fig. S2A**), and to the autologous tier 2 strain (BG505/T332N) (**Fig. 2B**). While 14/15 vaccinees mounted tier 1 neutralizing antibodies responses after the second or third dose, those receiving the higher 5µg 3M-052-AF/Alum tended to have higher autologous neutralizing titers. Moreover, in contrast to a previous report showing no autologous antibody neutralization after immunization with the closely related BG505 DS-SOSIP.554 (Trimer 4571) with alum adjuvant (Houser et al., 2022), in this study serum antibodies induced by BG505 SOSIP.664 with 3M-052/AF/Alum recognizing the tier 2 strain were detected in five vaccine recipients, including 3 after dose 2, and 2 additional participants after dose 3 (**Fig. 2B, Table S5**). After the third dose, neutralizing antibody activities were remarkably strong in participants 137-148 and 137-185 (ID50 titer, 404 and 8647, respectively), suggesting that three doses may be optimal with vaccine regimens using soluble trimer alone.

These data are the first demonstration that high titer tier 2 serum neutralizing responses can be elicited in humans by vaccination, albeit against the vaccine-matched strain. Antibodies that neutralize only the vaccine strain are unlikely to prevent HIV acquisition in humans following exposure to diverse circulating HIV strains, as has been shown in nonhuman primate SHIV challenge models (Pauthner et al., 2019; Charles et al., 2021). However, because heterologous tier 2 neutralizing antibodies have been reported following BG505 trimer immunization in cows (Sok et al., 2017), serum from donors generating autologous nAbs were also evaluated for neutralization against a representative panel of heterologous tier 2 viruses (see methods). No heterologous neutralizing antibody activity was observed.

This striking tier 2 neutralization was likely related to the use of the 3M-052-AF adjuvant. However, it is important to note the BG505 SOSIP.664 in this study and the BG505 DS-SOSIP.664 (Trimer 4571) in study VRC 018 differ by two amino acids designed to stabilize the trimeric structure (Kwon et al., 2015). Therefore, we cannot determine with certainty that adjuvant alone was the contributing factor to the discrepancy in observed neutralizing activity. The pseudovirus-based assay used for measurement of neutralization was the same, so we believe that technical variation is unlikely to be a contributor.

We were struck by the heterogeneity in neutralizing responses among the participants we observed. This is reminiscent of NHP models, where approximately half of vaccinated animals do not develop autologous neutralizing titers even after three or four doses of BG505 SOSIP.664 gp140 immunizations (Antanasijevic et al., 2021). The underlying cause of heterogeneity in the neutralizing response is unclear but may be related to variations in the innate or adaptive immune responses. We therefore determined the magnitude of the circulating memory B cells binding the fluorescent trimer probe response. Trimer-binding IgG+ B cells were easily detectable after the 2^nd^ dose and frequencies were remarkably high after the 3^rd^ dose (**Fig. 2C**). However, no major differences in the trimer-speciic B cell frequencies were observed in between donors with and without autologous neutralizing antibodies, suggesting that the B cell response in peripheral blood is unlikely to strictly delineate neutralization.

### Frequency and functional cluster analysis of T cell responses

CD4+ T cell help can also influence the development of serum neutralization (Lee et al., 2020) and different cytokine profiles can influence the quality of T cell help provided to B cells (Künzli and Masopust, 2023). Thus, we evaluated trimer-specific peripheral blood CD4+ T cell cytokine expression following ex vivo stimulation with 15-mer peptides spanning the BG505 gp120 and gp41 ectodomain sequences. Of the measured cytokines, moderate IFN-C and/or IL-2 response rates were observed after the first dose (80-87.5%), and higher frequencies were observed after the second and third third doses (80-90%) (**Fig. 3A**). Low CD4+ T cell responses rates (<38%) and magnitude (median, <0.075% of CD4+ T cells expressing IFN-C and/or IL-2 to the BG505 gp41 ectodomain were observed after the third dose. Neither CD4+ Th2 cells, defined as cells expressing IL-4, IL-5, and/or IL-13, nor CD8+ T cell responses were detected after any dose.

**Figure 3.**
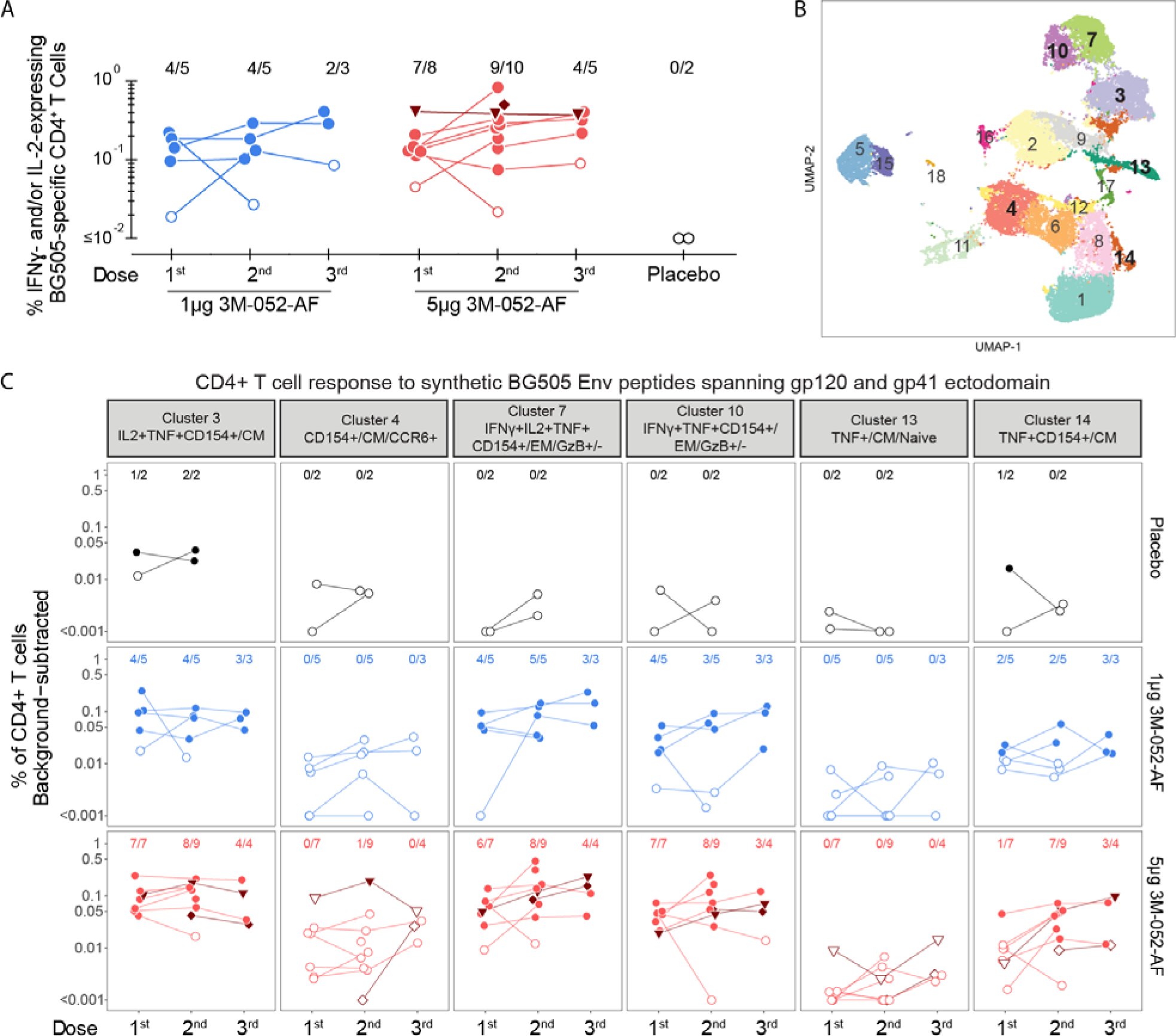
Antigen-specific CD4+ T cell responses elicited by BG505 SOSIP.664 gp140 with either 1µg or 5µg of 3M-052-AF + Alum. (A) Percent of CD4+ T cells expressing IFNγ and/or IL-2 in response to synthetic BG505 Env peptides spanning the gp120 and gp41 ectodomain using an ex vivo intracellular cytokine staining assay. Positive responses are shown with filled symbols. (B) Cluster analysis of CD4+ T cells from all treatment samples were combined. CD4+ T cells expressing at least 1 of 7 of the functional markers (IFNγ, IL-2, TNF, CD154, IL-4, IL-5/13, and IL-17a) were extracted and used as input. 18 clusters were identified. (C) Data for 6 clusters with positive responses (see Methods) were graphed as background-subtracted percentages of CD4+ T cells for the total Env response (gp120 and gp41). The dark red lines indicate CD4+ T cell responses in the two participants with the highest neutralizing titers (Figure 2B) after the third dose.

To better understand the CD4+ T cell phenotypic and functional profiles, we applied high-dimensional clustering analysis based on the markers expressed in the 27-color flow cytometry panel. Antigen-specific CD4+ T cell clusters emerged suggestive of a polyfunctional T cell phenotype (defined as cells expressing 3 or more critical markers, including IFN-C, IL-2, TNF, or CD40L) with minimal changes between the second and third dose. These findings support our initial observations that a relatively restricted blood CD4+ T cell phenotype was induced, and this phenotype did not change substantially between the second and third dose (**Figs. 3B, 3C**). The clusters were mixed between a central memory phenotype (Clusters 3, 4, 13, 14) and granzyme B expressing effector memory phenotype (Clusters 7 and 10). Overall, we observed no differences in the magnitude of T cell response profiles in participants who did or did not develop neutralizing antibodies.

### Strong neutralization associated with antibody response to the Env trimer apex

As there were no obvious differences in the antigen-specific T or B cell responses in participants who did or did not develop neutralization, we explored the possibility that epitope recognition sites of the antibodies mediating autologous tier 2 neutralization may be distinct from those for antibodies lacking this activity. We were intrigued that only base-directed responses were observed by EMPEM in the VRC018 trial (Hauser et al., 2022). We therefore hypothesized that 3M-052-AF could be providing a stimulus sufficient to overcome the immunodominant gp140 trimer base, which could lead to expansion of less frequent B cell clones capable of generating bnAb responses to the trimer apex. To test this hypothesis, we performed EMPEM analysis (**Fig. S1 and Fig. 4**). Similar to findings using the BG505 DS-SOSIP.664 trimer with Alum (Houser et al., 2022), we observed that 14/15 vaccinees mounted serum antibodies to the gp41 (ectodomain) base of the trimer after both the second and third doses. The discrepancy in the EMPEM results with the gp41 antibodies measured using the binding multiplex assay may relate to differences in the gp41 epitopes displayed in the BG505 SOSIP.664 trimer compared to the gp41 proteins included in the standard panel. In contrast to the VRC018 trial using Alum as an adjuvant, we observed responses directed at non-base epitopes located at the trimer apex. Most participants with detectable autologous neutralizing antibodies and both participants with serum neutralization ID50 titers >400 developed a response to the C3/V5 region of gp120 **(Fig. 4B and C)**. Only two participants developed responses to the gp120 glycan hole, one of whom had detectable serum autologous neutralizing activity after the third dose (Participant 9) (**Fig. 4C**). Additionally, the two participants with the highest tier 2 neutralizing titers also developed detectable protomers (**Fig. 4C**), which indicates antibodies capable of destabilizing the trimer base (Turner et al., 2021). We observed that participants receiving the 3M-052 5µg dose generally exhibited responses to more epitopes than participants receiving the 1µg dose after either the second or third dose.

**Figure 4.**
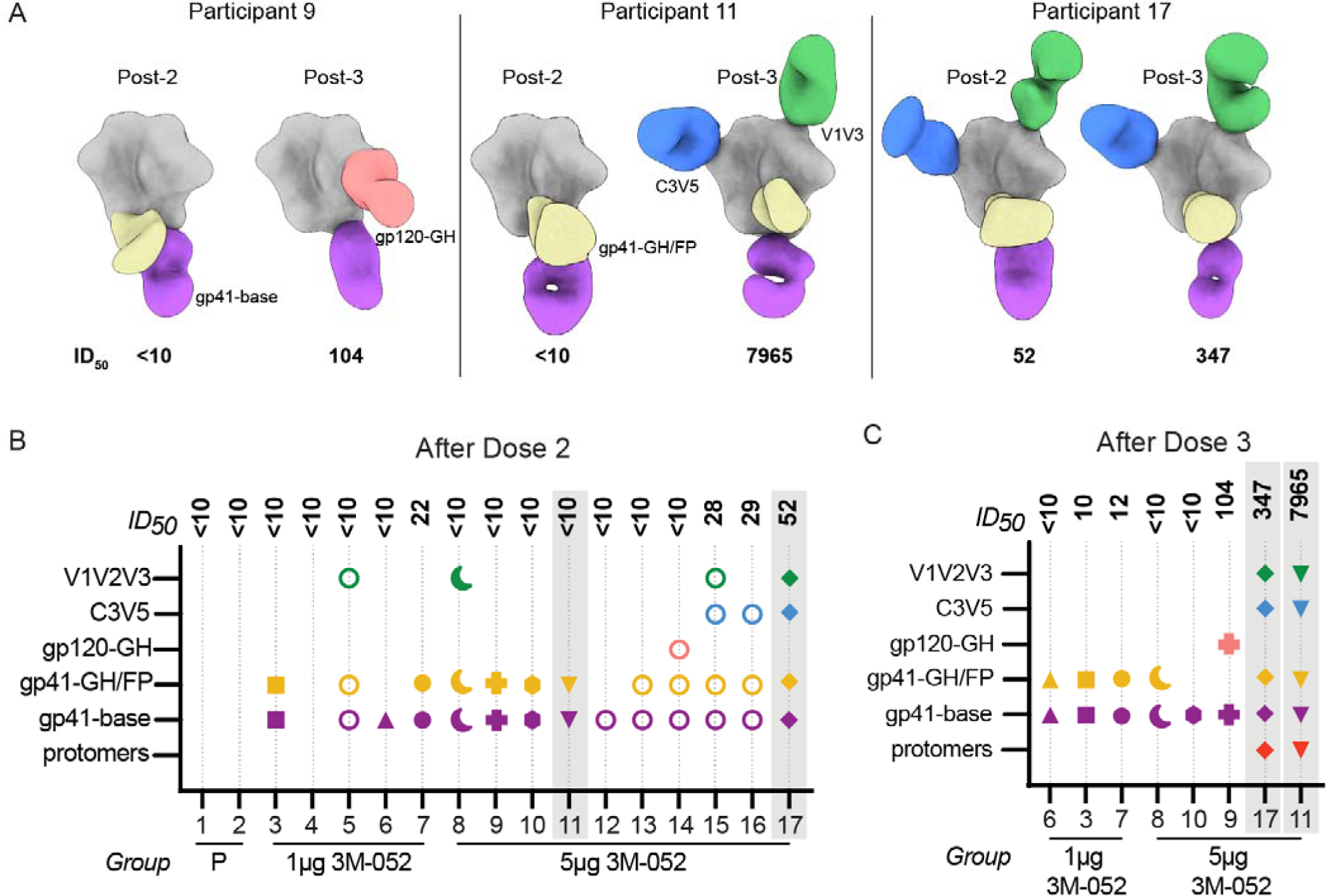
Three vaccine doses induce serum antibodies recognizing a wider range of Env epitopes, including V1/V2/V3 regions in selected participants with autologous neutralizing antibodies. (A) Representative EMPEM analysis of participants after the second and third doses. (B and C) Epitope regions recognized by polyclonal serum using negative stain electron microscopy after the 2^nd^ dose (B) and 3^rd^ dose. ID_50_ neutralizing titers to BG505/T332N are listed at the top. The X axis indicates the participant numbers and dose of 3M-052-AF. Colors correspond to epitope regions illustrated in A. Grey shading indicates the two individuals with the highest neutralizing titers after the third dose (see Figure 2).

Overall, BG505 SOSIP.664 formulated with 3M-052-AF + Alum led to an antibody response directed at the apex of the trimer. However, it was unclear whether the apex-directed antibodies were capable of neutralization. If autologous neutralizing antibodies are directed against other, more variable epitopes specific to the vaccine strain, it would suggest limited potential for B cell lineages capable of producing autologous neutralizing antibodies to evolve as lineages capable of producing bnAbs. To determine if the antibodies targeting epitopes at the trimer apex were responsible for neutralization, we introduced mutations to the BG505 pseudovirus to disrupt antibody binding in the neutralization assay. One specific set of mutations in BG505/T332N.133aN.136a are in the V1/V2/V3 region (**Fig. 5A**). When these mutations were introduced, we observed a roughly 50% reduction in the serum neutralization activity in participants 11 and 17 showing the highest neutralizing titer (**Figs. 5B and S2**), demonstrating that this epitope substantially contributes to the observed serum neutralization activity. Conversely, no reduction in serum antibody neutralization was observed in pseudoviruses with mutations at T465N (C3V5 region), I358T (C3V5 region), or S241N/P291T (gp120 glycan hole), suggesting that although these epitopes were recognized by polyclonal serum in participants with neutralizing activity, antibodies directed to these regions were not directly responsible for the overall neutralizing activity. Both mutations that lead to a reduction in neutralization activity are in regions of HIV Env that have substantial variation in clinical isolates.

**Figure 5.**
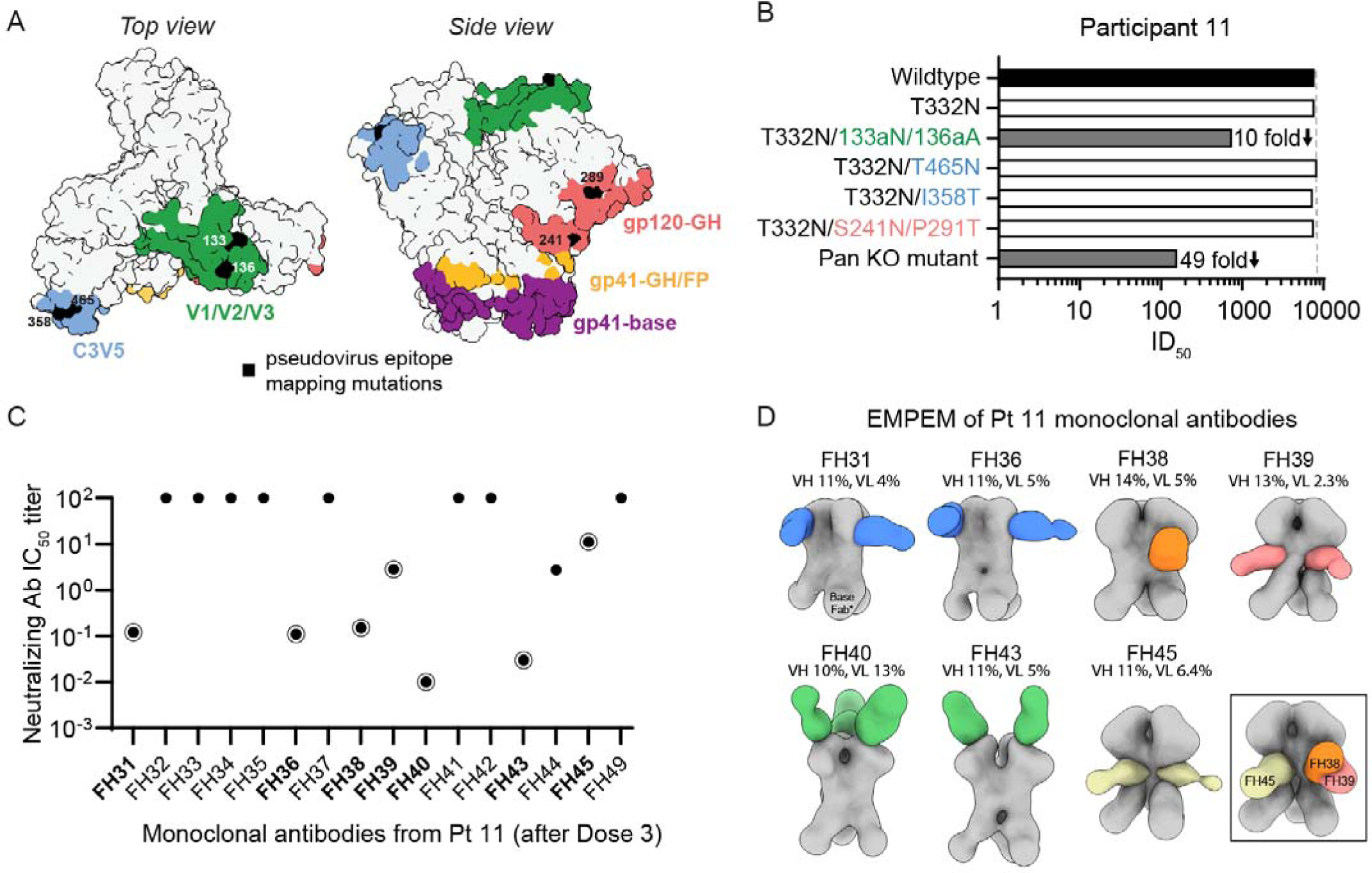
Mapping BG505 SOSIP.664 gp140 epitope regions recognized by serum and monoclonal neutralizing antibody responses. (A) Schematic representation of the epitope surface targeted by polyclonal serum antibodies to BG505 SOSIP.664 gp140, as determined by published cryo-EM structures (Antanasijevic *et al*., 2021). The location of mutations used in pseudovirus-based epitope mapping are marked at indicated positions. (B) Serum antibody neutralization titers for participant 11 were mapped using pseudoviruses with mutations introduced at indicated positions and color coded as in A. “Pan KO mutant” includes the following mutations: T332N, T465N, S241N, 133aA, and 141aN. Open bars represent titers that were not impacted by the mutations (<three-fold reduction relative to wildtype BG505 virus). (C) IC50 of monoclonal antibodies against BG505/T332N isolated from Participant 11 after third dose. Circled points indicate samples also analyzed by EMPEM. (D) EMPEM of indicated monoclonal antibodies with % somatic hypermutation of heavy and light chains indicated.

Our observation that the introduced BG505 SOSIP.664 mutations did not completely abrogate neutralizing activity is consistent with findings using mutant antigenic profiling indicating that recognition of multiple epitopes may contribute to neutralization in a given polyclonal antibody response (Dingens et al., 2021). To confirm our observations made with pseudovirus mutants, we isolated BG505-specific B cells from Participant 11 with the highest neutralization activity and expressed the isolated B cell receptors as a series of monoclonal antibodies, with clonotype size and sequence features described in **Table S6**. Among the 16 antibodies binding BG505 SOSIP.664 **(Fig. S3A),** 8 isolated antibodies had detectable neutralizing activities (Fig. 5C). Only one expressed antibody (FH38) had low levels of neutralizing titers against two out of 11 heterologous tier 2 viruses **(Fig. S3B).** When the epitopes of these expressed antibodies were assessed by EMPEM, we determined that the mAbs with the most potent neutralization activity (FH40 and FH43) targeted the C3/V5 region **(Fig. 5D)**. Antibodies targeting the C3/V5 region were only observed in participants with autologous neutralizing serum, supporting observations from vaccinated macaques that this region could be a source of autologous bnAb in humans (Klasse et al., 2018; Zhao et al., 2020; Charles et al., 2021). Interestingly, unlike macaques immunized with an analogous BG505 SOSIP MD39 trimer, only two participants developed detectable responses against the gp120 N241/N289 glycan hole using polyclonal serum (**Fig. 4B and C**). This epitope is highly specific to the BG505 SOSIP trimer, has been frequently observed in macaques immunized with BG505-derived trimers and may be less desirable due to its absence in most wild-type HIV-1 strains (McCoy et al., 2016; Antanasijevic et al., 2021). Consistent with previous observations in rabbits, the two participants 11 and 17 who developed autologous neutralizing antibodies also developed antibodies capable of reacting to protomers, indicative of antibody-dependent trimer degradation (Turner et al., 2021).

### Summary

Forty years of research has failed to generate a licensed HIV vaccine. Historically, the approach to developing new adjuvant formulations for clinical use has been profoundly slow (Gregorio et al., 2008). To accelerate process toward an effective HIV vaccine, this study was designed to introduce the native-like Env trimer into humans, particularly since stabilized trimers may have utility as a booster in sequential vaccine regimens designed to induce bnAbs. Simultaneously, we were able to evaluate in humans a new adjuvant formulation that has induced strong Tfh and germinal center responses as well as persistent long-lived plasma cells in nonhuman primates (Kasturi et al., 2020). We report that the BG505 SOSIP.664 trimer with 3M-052-AF/alum can be safely administered in humans and that antibody responses generated can neutralize a tier 2 circulating viral strain.

Our findings strongly suggest that an effective adjuvant is likely to play a role in improving potency of the neutralizing response to HIV immunogens. High-titer autologous neutralizing antibody responses were only observed in the 5µg dose group. Whether a lower dose would be appropriate as an adjuvant for vaccines targeting different antigens is also unclear as HIV presents many unique challenges; only a minority of people living with HIV develop bnAbs, in contrast to the near universal neutralization observed after SARS-CoV-2 vaccination and acquisition.

Comparison with published results using a virtually identical trimer adjuvanted with Alum strongly suggest that a more potent adjuvant can expand the range of epitopes recognized beyond the more restricted, immunodominant epitopes at the trimer base. Importantly, diversifying the antibodies beyond the base recognition directly mediated serum neutralization. The areas associated with neutralization are not known bnAb epitopes and mutations in these areas are relatively well tolerated HIV-1. Although these observations suggest that the unmodified BG505 SOSIP.664 is unlikely to directly protect against HIV, we note that antibodies with HIV tier 2-3 neutralization has not been observed in humans previously (even with closely related vaccine regimens). Autologous tier 2 neutralizing activities may represent a threshold response required for all bnAb-based vaccine strategies, and future studies will be necessary to confirm this.

There are several limitations of our study, including variable timing between administration of vaccinations as a result of the COVID pandemic and the small sample size. Analyses of the durability of the immune response driven by the 3M-052-AF + Alum adjuvant and how this compares to other adjuvants is in progress (HVTN 137 Part B). Our results suggest that additional vaccine strategies beyond recombinant trimer designs of wild-type Env antigens are likely to be required to elicit the broader serum neutralizing antibody response necessary to prevent HIV acquisition, but that the choice of adjuvant may influence the success based upon recombinant protein approaches. The first clinical experience with 3M-052-AF/Alum as a vaccine adjuvant reported here provides a reassuring safety outcome and a promising immune response that support further clinical testing of this adjuvant.

### Conflict of Interest

NR is a safety clinical trial consultant for ICON and EMMES, serves on the advisory boards for Sanofi, Seqirus and Moderna, and Emory receives funds for NR to conduct research from Sanofi, Lilly, Merck, Quidel and Pfizer. MT is a contract worker for 3M. Fred Hutch Cancer Center receives funds for MJM to conduct research from Janssen, Sanofi, Regeneron and Moderna.

## Acknowledgments and funding

We thank all investigators and staff at the clinical trial sites and the study volunteers for completing this complex study, particularly during the SARS-CoV-2 pandemic. We appreciate the helpful discussions with John P Moore at Cornell-Weill and Rogier Sanders at the University of Amsterdam for use of the BG505 SOSIP.664. We thank Michael Pensiero, Mary Marovich and Patricia D’Souza at the NIAID Division of AIDS for sponsorship and support of the clinical trial, and Thomas Hassell and Mark Feinberg at IAVI and Al Cupo at Weill Cornell Medicine for production and use of the trimer. We also thank Al Cupo, PJ Klasse (Weill Cornell Medicine) and Marit van Gils (Academic Universities Medicine Center) for the production, provision, and advice on the relevant analytical probes, including the tagged BG505 trimers for immune response analytics. We appreciate the contributions of Ian Wilson to the design and production of BG505.664 trimers. We thank Michael Pensiero, Mary Marovich and Patricia D’Souza at the NIAID Division of AIDS, and Thomas Hassell and Mark Feinberg at IAVI for production and use of the trimer. We thank Corey Casper and Christopher Fox at AAHI providing the 3M-052-AF adjuvant and Nina Russell and Susan Barnett and Pervin Anklesaria (BMGF) for guidance in advancing the adjuvant study. We thank the laboratory investigators and staff for their efforts, including Kristen Cohen at FHCC (now at Moderna; Sara Richey and Jonathan Torres for EMPEM studies (Scripps Research Institute); and the staff at the HVTN Laboratories and Operations, including John Hural, Jennifer Hanke at Fred Hutch, as well as Jack Heptinstall, Yong Lin, Kelly Seaton, Sheetal Sawant and Lu Zhang for technical and analytical expertise. We also thank Stephen Voght and Gabe Murphy for critical reading of the manuscript.

The trial was funded through National Institute of Allergy and Infectious Disease of the National Institutes of Health under Grants UM1 AI068614 [HVTN LOC], UM1 AI068635 [HVTN SDMC], and UM1 AI068618 [HVTN LC], UM1 AI069481 [Seattle CTU], and AI110657 HIVRAD [JPM]. Funding from the Gates Foundation OPP1107954 to Fred Hutchison (MJM) for the manufacture/formulation of 3M-052 AF, and nonclinical studies; OPP1147661 to IAVI for GMP manufacturing of the trimer protein supply for this study; INV-002022 [RWS] and INV-002916 to Scripps/Ward lab for early EMPEM analyses [ADW].

## Author contributions

WOH, MS, HJ, NR, AKD, ZS, NR and MJM designed the clinical protocol. MJM, JM, IF, MCK, LRB, NR, and recruited participants and generated clinical data. MT, AKD, ZS, MS, RWS, JPM and MJM generated clinical trial material. GO, HLT, and ABW generated and analyzed EMPEM data. GDT and DCM generated antibody data. SDR and MJM generated CD4 T cell data. KRP and MJM generated B cell data. HJ performed statistical and data analysis. WOH, KRP, MS, GO, HJ, AWD, CD, AKD, ZS, MCK, OH, ABW, GT, DM, NR, MJM drafted the initial manuscript. All authors provided guidance and helped with revisions and provided final approval for it to be published.

