## Supplemental Figures 1-3 and Supplemental Tables 1-5 for "HIV BG505 SOSIP.664 trimer with 3M-052-AF/alum induces human autologous tier-2 neutralizing antibodies"

**Supplementary Materials**

Supplemental Figure 1. Electron microscopy epitope mapping data of polyclonal antibodies in sera after the second and third immunizations.

Supplemental Figure 2. Neutralizing antibody responses to tier 1 MW965.26 and molecular mapping of neutralization activities in Participant 17.

Supplemental Figure 3. Monoclonal antibody binding and neutralization

Supplemental Table 1. Study participant demographics.

Supplemental Table 2. Visit schedule, participant PubID, and participant numbers.

Supplemental Table 3. Maximum local reactogenicity for all participants.

Supplemental Table 4. Maximum systemic reactogenicity for all participants.

Supplemental Table 5. Monoclonal antibody sequence information.

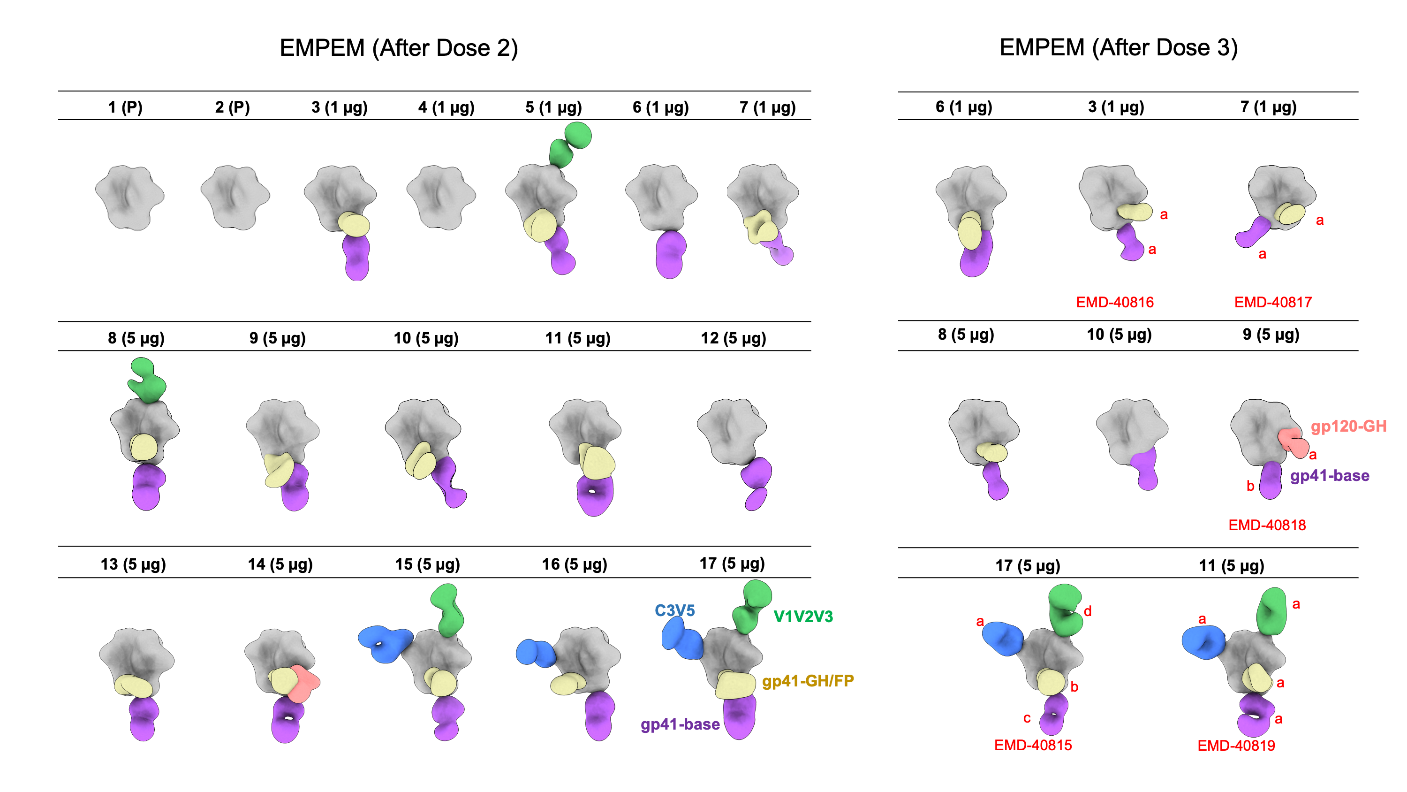

**Supplemental Figure 1. Electron microscopy serum polyclonal antibody epitope mapping data.** Segmented maps representing polyclonal antibody responses detected against BG505 SOSIP antigen for each participant after 2 or 3 doses. Participant numbers are the same as **Figure 4**. Treatment group denoted next to each participant number (P: placebo, 1 µg 3M-052, 5 µg 3M-052). Polyclonal epitopes are colored as defined for participant 17 (after dose 2) and participant 9 (after dose 3). Representative maps have been deposited to the Electron Microscopy Data Bank under the listed accession codes: antibodies with red letter “a” are found in the main EMDB entry map; antibodies with letters “b”, “c”, or “d” can be found as “additional maps” under the Download tab of the respective entry on <https://www.emdataresource.org/>.

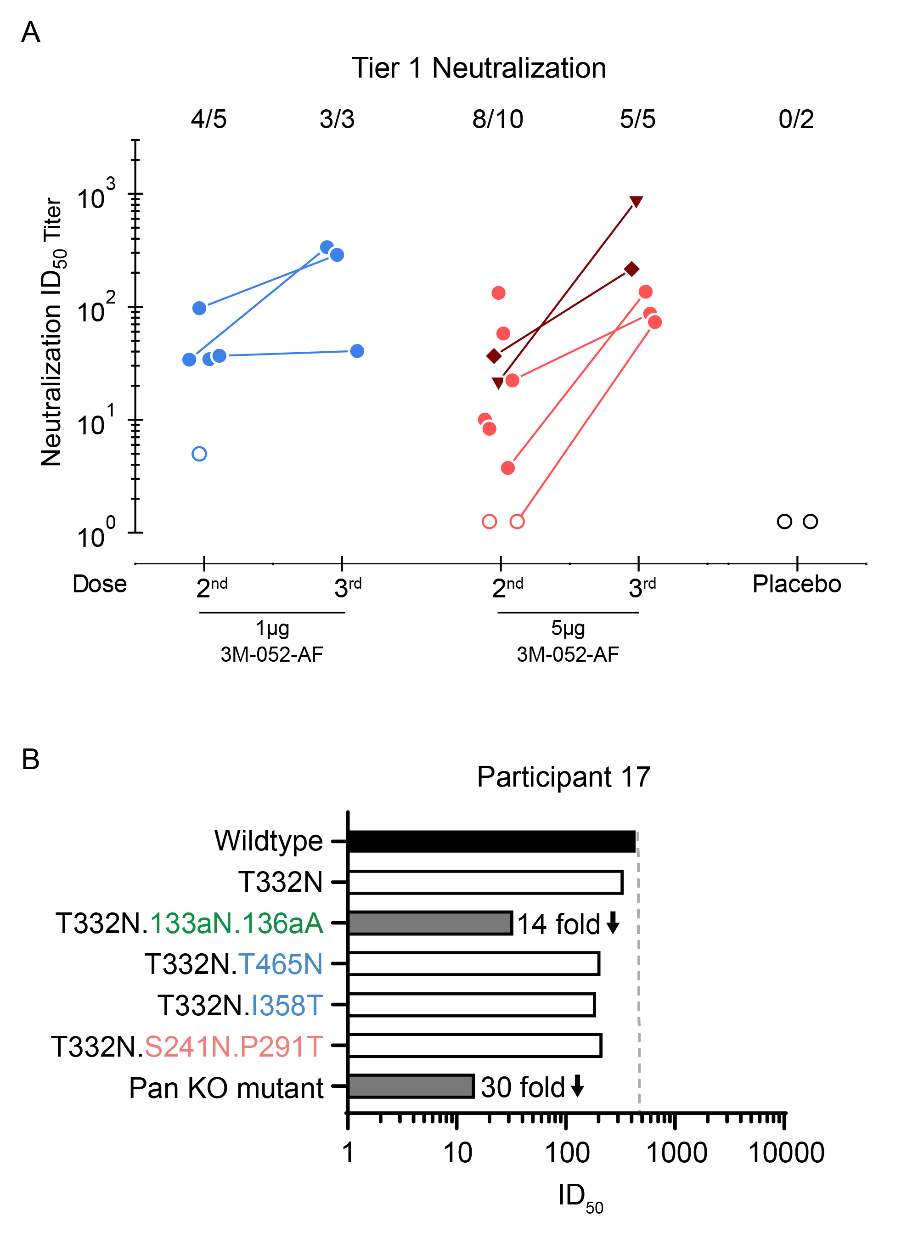

**Supplemental Figure 2. Antibody neutralization.** (A) ID_50_ Neutralizing antibody titers against the tier 1 virus MW965.26 using the TZMbl neutralization assay after the indicated vaccine dose. The participants indicated by darker shading are the same two individuals with the highest neutralizing titers after the third dose (participant 11 indicated by ▼ and Participant 17 indicated by ♦). (B) Pseudovirus neutralization titers for participant 17. Neutralization was mapped using mutant pseudoviruses with mutations introduced at indicated positions and color coded as in A. “Pan KO mutant” includes the following mutations: T332N, T465N, S241N, 133aA, and 141aN. Open bars represent titers that were not impacted by the mutations (<three-fold reduction relative to Wildtype virus).

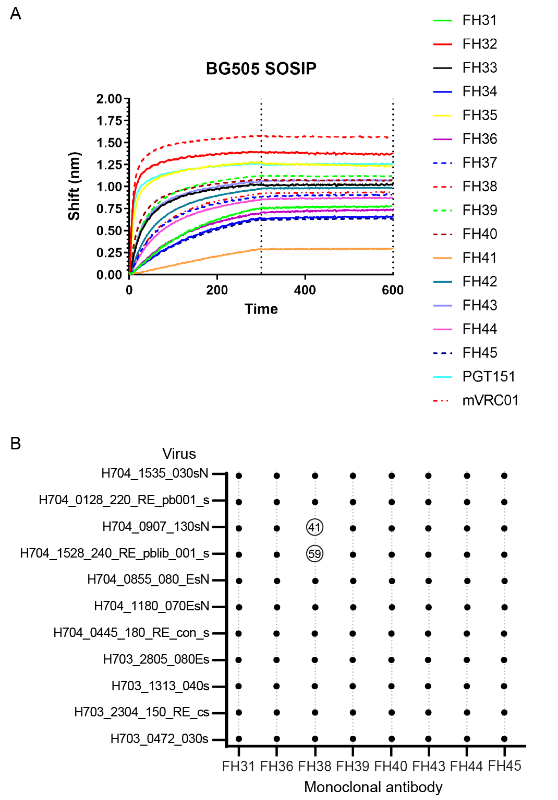

**Supplemental Figure 3. Monoclonal antibody binding and neutralization.** (A) Biolayer inferometry binding of isolated monoclonal antibodies from clonotypes of single B cells sorted and sequenced. VRC01 and PGT151 are monoclonals with known binding activity included as representative controls. (B) Pseudovirus neutralization titers against a virus panel selected based on tier 2 phenotype.

**Supplemental Table 1. Study participant demographics.**

|  | **1μg 3M-052** | **5μg 3M-052** | **Placebo** |
| --- | --- | --- | --- |
| Assigned Sex at Birth |  |  |  |
| Male | 1 (20%) | 4 (40%) | 0 |
| Female | 4 (80%) | 6 (60%) | 2 (100%) |
| Gender Identity |  |  |  |
| Gender Queer | 1 (20%) | 1 (10%) | 0 |
| Female | 2 (40%) | 5 (50%) | 2 (100%) |
| Male | 2 (40%) | 4 (40%) | 0 |
| Ethnicity |  |  |  |
| Hispanic or Latino/a | 0 | 1 (10%) | 1 (50%) |
| Not Hispanic or Latino/a | 5 (100%) | 9 (90%) | 1 (50%) |
| Race |  |  |  |
| White | 3 (60%) | 8 (80%) | 1 (50%) |
| Black or African American | 1 (20%) | 0 | 1 (50%) |
| Asian | 1 (20%) | 2 (20%) | 0 |
| Age (Years) |  |  |  |
| 18 - 20 | 0 | 1 (10%) | 0 |
| 21 - 30 | 4 (80%) | 7 (70%) | 2 (100%) |
| 31 - 40 | 1 (20%) | 2 (20%) | 0 |
| Median | 28 | 26 | 24 |
| Range | 21-35 | 20-34 | 23-24 |

**Supplemental Table 2.** Visit schedule, participant PubID, and participant numbers.

| **Participant** | **PubID** | **Visit 2** | **Visit 6** | **Visit 7** | **Visit 9** | **Visit 11** | **Visit 13** | **Visit 14** | **Last Contact** |
| --- | --- | --- | --- | --- | --- | --- | --- | --- | --- |
|  |  | **Dose 1** |  | **Dose 2** |  | **Dose 3^** |  |  |  |
| 1 | 137-115 | Day 0 | Day 10 | Day 90 | Day 105 |  |  | Day 212 | Day 406 |
| 2 | 137-239 | Day 0 | Day 10 | Day 52 | Day 66 |  |  | Day 278 | Day 440 |
| 3 | 137-030 | Day 0 | Day 10 | Day 210 | Day 225 | Day 498 | Day 519 |  | Day 883 |
| 4 | 137-206 | Day 0 | Day 14 | Day 54 | Day 75 |  |  | Day 229 | Day 415 |
| 5 | 137-247 | Day 0 | Day 14 | Day 223 | Day 237 |  |  | Day 279 | Day 494 |
| 6 | 137-109 | Day 0 | Day 15 | Day 50 | Day 62 | Day 462 | Day 477 |  | Day 843 |
| 7 | 137-052 | Day 0 | Day 14 | Day 146 | Day 160 | Day 483 | Day 497 |  | Day 869 |
| 8 | 137-003 | Day 0 | Day 13 | Day 50 | Day 62 | Day 304 | Day 317 |  | Day 673 |
| 9 | 137-187 | Day 0 | Day 13 | Day 55 | Day 69 | Day 403 | Day 447 |  | Day 818 |
| 10 | 137-214 | Day 0 | Day 15 | Day 57 | Day 75 | Day 399 | Day 411 |  | Day 806 |
| 11 | 137-185 | Day 0 | Day 12 | Day 49 | Day 61 | Day 368 | Day 382 |  | Day 733 |
| 12 | 137-025 | Day 0 | Day 14 | Day 137 | Day 154 |  |  | Day 244 | Day 244 |
| 13 | 137-018 | Day 0 | Day 14 | Day 49 | Day 62 | Day 364 | * |  | Day 728 |
| 14 | 137-116 | Day 0 | Day 14 | Day 54 | Day 70 |  |  | Day 196 | Day 401 |
| 15 | 137-170 | Day 0 | Day 17 | * | * |  |  | Day 240 | Day 240 |
| 16 | 137-165 | Day 0 | Day 14 | Day 48 | Day 81 |  |  |  | Day 130 |
| 17 | 137-148 | Day 0 | Day 20 | Day 126 | Day 139 | Day 492 | Day 504 |  | Day 678 |

^optional third dose

*denotes missed visit

Date of last contact is defined as the date of the last scheduled visit, which was an AESI health contact at Month 14 or Visit 16 at Month 18, if the participant received the third dose.  If a participant terminated early, their last contact is their last contact with the site or their last study visit, whichever was more recent.

Visits 11 and 13 were only scheduled for participants who received the third dose. Visit 14 was a scheduled visit for those who only received two doses.

**Supplemental Table 3.** Maximum local reactogenicity for all participants.

|  | **1μg 3M-052** | | | | **5μg 3M-052** | | | | **Placebo** | | |
| --- | --- | --- | --- | --- | --- | --- | --- | --- | --- | --- | --- |
| **Symptom / Severity** | **Vacc 1**  **(N=5)** | **Vacc 2**  **(N=4)** | **Vacc 3**  **(N=3)** | **All Vacc (N=5)** | **Vacc 1 (N=10)** | **Vacc 2**  **(N=9)** | **Vacc 3**  **(N=6)** | **All Vacc (N=10)** | **Vacc 1**  **(N=2)** | **Vacc 2**  **(N=2)** | **All Vacc (N=2)** |
| **Participants with 1 or more local**  **reactogenicities** None | 0 | 1 (25%) | 0 | 0 | 0 | 0 | 1 (16.7%) | 0 | 1 (50%) | 2 (100%) | 1 (50%) |
| Not gradable | 0 | 0 | 0 | 0 | 0 | 0 | 0 | 0 | 0 | 0 | 0 |
| Grade 1: Mild | 4 (80%) | 3 (75%) | 3 (100%) | 4 (80%) | 4 (40%) | 6 (66.7%) | 3 (50%) | 3 (30%) | 1 (50%) | 0 | 1 (50%) |
| Grade 2: Moderate | 1 (20%) | 0 | 0 | 1 (20%) | 4 (40%) | 3 (33.3%) | 2 (33.3%) | 5 (50%) | 0 | 0 | 0 |
| Grade 3: Severe | 0 | 0 | 0 | 0 | 2 (20%) | 0 | 0 | 2 (20%) | 0 | 0 | 0 |
| Grade 4: Complications, Potentially life-threatening | 0 | 0 | 0 | 0 | 0 | 0 | 0 | 0 | 0 | 0 | 0 |
| **Erythema/Redness** None | 4 (80%) | 2 (50%) | 3 (100%) | 3 (60%) | 7 (70%) | 8 (88.9%) | 5 (83.3%) | 7 (70%) | 2 (100%) | 2 (100%) | 2 (100%) |
| Not gradable | 1 (20%) | 2 (50%) | 0 | 2 (40%) | 1 (10%) | 1 (11.1%) | 0 | 1 (10%) | 0 | 0 | 0 |
| Grade 1: 2.5 - <5cm dim. | 0 | 0 | 0 | 0 | 0 | 0 | 0 | 0 | 0 | 0 | 0 |
| Grade 2: 5 - <10cm dim. | 0 | 0 | 0 | 0 | 0 | 0 | 1 (16.7%) | 0 | 0 | 0 | 0 |
| Grade 3: >= 10cm dim. | 0 | 0 | 0 | 0 | 2 (20%) | 0 | 0 | 2 (20%) | 0 | 0 | 0 |
| Grade 3: complications AE | 0 | 0 | 0 | 0 | 0 | 0 | 0 | 0 | 0 | 0 | 0 |
| Grade 4: complications AE | 0 | 0 | 0 | 0 | 0 | 0 | 0 | 0 | 0 | 0 | 0 |
| **Induration/Swelling** None | 4 (80%) | 4 (100%) | 3 (100%) | 4 (80%) | 7 (70%) | 8 (88.9%) | 6 (100%) | 7 (70%) | 2 (100%) | 2 (100%) | 2 (100%) |
| Not gradable | 0 | 0 | 0 | 0 | 1 (10%) | 1 (11.1%) | 0 | 1 (10%) | 0 | 0 | 0 |
| Grade 1: 2.5 - <5cm diam. | 0 | 0 | 0 | 0 | 0 | 0 | 0 | 0 | 0 | 0 | 0 |
| Grade 2: 5 - <10cm diam. | 1 (20%) | 0 | 0 | 1 (20%) | 0 | 0 | 0 | 0 | 0 | 0 | 0 |
| Grade 3: >= 10cm diam. | 0 | 0 | 0 | 0 | 2 (20%) | 0 | 0 | 2 (20%) | 0 | 0 | 0 |
| Grade 3: complications AE | 0 | 0 | 0 | 0 | 0 | 0 | 0 | 0 | 0 | 0 | 0 |
| Grade 4: complications AE | 0 | 0 | 0 | 0 | 0 | 0 | 0 | 0 | 0 | 0 | 0 |

*Supplemental Table 3 continued*

|  | 1μg 3M-052 | | | | 5μg 3M-052 | | | | Placebo | | |
| --- | --- | --- | --- | --- | --- | --- | --- | --- | --- | --- | --- |
| Symptom / Severity | Vacc 1  (N=5) | Vacc 2  (N=4) | Vacc 3  (N=3) | All Vacc (N=5) | Vacc 1 (N=10) | Vacc 2  (N=9) | Vacc 3  (N=6) | All Vacc (N=10) | Vacc 1  (N=2) | Vacc 2  (N=2) | All Vacc (N=2) |
| **Erythema and/or Induration**  None | 3 (60%) | 2 (50%) | 3 (100%) | 2 (40%) | 7 (70%) | 8 (88.9%) | 5 (83.3%) | 7 (70%) | 2 (100%) | 2 (100%) | 2 (100%) |
| Not gradable | 1 (20%) | 2 (50%) | 0 | 2 (40%) | 1 (10%) | 1 (11.1%) | 0 | 1 (10%) | 0 | 0 | 0 |
| Grade 1: 2.5 - <5cm dim. | 0 | 0 | 0 | 0 | 0 | 0 | 0 | 0 | 0 | 0 | 0 |
| Grade 2: 5 - <10cm dim. | 1 (20%) | 0 | 0 | 1 (20%) | 0 | 0 | 1 (16.7%) | 0 | 0 | 0 | 0 |
| Grade 3: >= 10cm dim. | 0 | 0 | 0 | 0 | 2 (20%) | 0 | 0 | 2 (20%) | 0 | 0 | 0 |
| Grade 3: complications AE | 0 | 0 | 0 | 0 | 0 | 0 | 0 | 0 | 0 | 0 | 0 |
| Grade 4: complications AE | 0 | 0 | 0 | 0 | 0 | 0 | 0 | 0 | 0 | 0 | 0 |
| **Pain and/or Tenderness** None | 0 | 1 (25%) | 0 | 0 | 0 | 0 | 1 (16.7%) | 0 | 1 (50%) | 2 (100%) | 1 (50%) |
| Mild | 4 (80%) | 3 (75%) | 3 (100%) | 4 (80%) | 5 (50%) | 6 (66.7%) | 4 (66.7%) | 4 (40%) | 1 (50%) | 0 | 1 (50%) |
| Moderate | 1 (20%) | 0 | 0 | 1 (20%) | 5 (50%) | 3 (33.3%) | 1 (16.7%) | 6 (60%) | 0 | 0 | 0 |
| Severe | 0 | 0 | 0 | 0 | 0 | 0 | 0 | 0 | 0 | 0 | 0 |
| Potentially life-threatening | 0 | 0 | 0 | 0 | 0 | 0 | 0 | 0 | 0 | 0 | 0 |

For a given sign or symptom, each participant's reactogenicity is counted once under the maximum severity for all post-administration assessments at study product administration visits. Pain, tenderness, erythema, and induration symptoms are considered local reactogenicity if onset was within 7 days following study product administration. The item Erythema and/or Induration is the maximum of the individual variables Erythema and Induration.

Reactogenicities are graded according to DAIDS AE Grading Table Version 2.1, July 2017.

1μg 3M-052: BG505 SOSIP.664 gp140 & (1 mcg 3M-052-AF + Alum) month (0,2)

5μg 3M-052: BG505 SOSIP.664 gp140 & (5 mcg 3M-052-AF + Alum) month (0,2)

Placebo for BG505 SOSIP.664 gp140 & adjuvant month (0,2)

**Supplemental Table 4.** Maximum systemic reactogenicity for all participants.

|  | 1μg 3M-052 | | | | 5μg 3M-052 | | | | Placebo | | |
| --- | --- | --- | --- | --- | --- | --- | --- | --- | --- | --- | --- |
| Symptom / Severity | Vacc 1  (N=5) | Vacc 2  (N=4) | Vacc 3  (N=3) | All Vacc (N=5) | Vacc 1 (N=10) | Vacc 2  (N=9) | Vacc 3  (N=6) | All Vacc (N=10) | Vacc 1  (N=2) | Vacc 2  (N=2) | All Vacc (N=2) |
| **Participants with 1 or more**  **systemic reactogenicities** None | 0 | 0 | 0 | 0 ( 0%) | 0 ( 0%) | 1 (11.1%) | 2 (33.3%) | 0 | 2 (100%) | 2 (100%) | 2 (100%) |
| Mild | 2 (40%) | 1 (25%) | 0 | 1 (20%) | 2 (20%) | 2 (22.2%) | 1 (16.7%) | 2 (20%) | 0 | 0 | 0 |
| Moderate | 3 (60%) | 3 (75%) | 2 (66.7%) | 3 (60%) | 6 (60%) | 5 (55.6%) | 1 (16.7%) | 5 (50%) | 0 | 0 | 0 |
| Severe | 0 | 0 | 1 (33.3%) | 1 (20%) | 2 (20%) | 1 (11.1%) | 2 (33.3%) | 3 (30%) | 0 | 0 | 0 |
| Potentially Life-Threatening | 0 | 0 | 0 | 0 ( 0%) | 0 | 0 | 0 | 0 | 0 | 0 | 0 |
| **Malaise and/or Fatigue** None | 1 (20%) | 1 (25%) | 0 | 0 | 1 (10%) | 1 (11.1%) | 2 (33.3%) | 0 | 2 (100%) | 2 (100%) | 2 (100%) |
| Mild | 2 (40%) | 1 (25%) | 1 (33.3%) | 2 (40%) | 1 (10%) | 4 (44.4%) | 1 (16.7%) | 2 (20%) | 0 | 0 | 0 |
| Moderate | 2 (40%) | 2 (50%) | 1 (33.3%) | 2 (40%) | 6 (60%) | 3 (33.3%) | 2 (33.3%) | 5 (50%) | 0 | 0 | 0 |
| Severe | 0 | 0 | 1 (33.3%) | 1 (20%) | 2 (20%) | 1 (11.1%) | 1 (16.7%) | 3 (30%) | 0 | 0 | 0 |
| Potentially Life-Threatening | 0 | 0 | 0 | 0 | 0 | 0 | 0 | 0 | 0 | 0 | 0 |
| **Myalgia** None | 2 (40%) | 4 (100%) | 0 | 1 (20%) | 1 (10%) | 3 (33.3%) | 3 (50%) | 1 (10%) | 2 (100%) | 2 (100%) | 2 (100%) |
| Mild | 1 (20%) | 0 | 2 (66.7%) | 1 (20%) | 3 (30%) | 2 (22.2%) | 3 (50%) | 2 (20%) | 0 | 0 | 0 |
| Moderate | 2 (40%) | 0 | 0 | 2 (40%) | 5 (50%) | 3 (33.3%) | 0 | 6 (60%) | 0 | 0 | 0 |
| Severe | 0 | 0 | 1 (33.3%) | 1 (20%) | 1 (10%) | 1 (11.1%) | 0 | 1 (10%) | 0 | 0 | 0 |
| Potentially Life-Threatening | 0 | 0 | 0 | 0 | 0 | 0 | 0 | 0 | 0 | 0 | 0 |

*Supplemental Table 4 continued*

|  | 1μg 3M-052 | | | | 5μg 3M-052 | | | | Placebo | | |
| --- | --- | --- | --- | --- | --- | --- | --- | --- | --- | --- | --- |
| Symptom / Severity | Vacc 1  (N=5) | Vacc 2  (N=4) | Vacc 3  (N=3) | All Vacc (N=5) | Vacc 1 (N=10) | Vacc 2  (N=9) | Vacc 3  (N=6) | All Vacc (N=10) | Vacc 1  (N=2) | Vacc 2  (N=2) | All Vacc (N=2) |
| **Headache** None | 1 (20%) | 1 (25%) | 0 | 0 | 2 (20%) | 2 (22.2%) | 3 (50%) | 0 | 2 (100%) | 2 (100%) | 2 (100%) |
| Mild | 2 (40%) | 2 (50%) | 2 (66.7%) | 3 (60%) | 2 (20%) | 4 (44.4%) | 2 (33.3%) | 4 (40%) | 0 | 0 | 0 |
| Moderate | 2 (40%) | 1 (25%) | 1 (33.3%) | 2 (40%) | 6 (60%) | 3 (33.3%) | 0 | 5 (50%) | 0 | 0 | 0 |
| Severe | 0 | 0 | 0 | 0 | 0 | 0 | 1 (16.7%) | 1 (10%) | 0 | 0 | 0 |
| Potentially Life-Threatening | 0 | 0 | 0 | 0 | 0 | 0 | 0 | 0 | 0 | 0 | 0 |
| **Nausea** None | 5 (100%) | 3 (75%) | 2 (66.7%) | 3 (60%) | 7 (70%) | 5 (55.6%) | 5 (83.3%) | 3 (30%) | 2 (100%) | 2 (100%) | 2 (100%) |
| Mild | 0 | 1 (25%) | 1 (33.3%) | 2 (40%) | 2 (20%) | 2 (22.2%) | 1 (16.7%) | 5 (50%) | 0 | 0 | 0 |
| Moderate | 0 | 0 | 0 | 0 | 1 (10%) | 2 (22.2%) | 0 | 2 (20%) | 0 | 0 | 0 |
| Severe | 0 | 0 | 0 | 0 | 0 | 0 | 0 | 0 | 0 | 0 | 0 |
| Potentially Life-Threatening | 0 | 0 | 0 | 0 | 0 | 0 | 0 | 0 | 0 | 0 | 0 |
| **Chills**  None | 3 (60%) | 3 (75%) | 0 | 2 (40%) | 4 (40%) | 5 (55.6%) | 4 (66.7%) | 4 (40%) | 2 (100%) | 2 (100%) | 2 (100%) |
| Mild | 1 (20%) | 1 (25%) | 1 (33.3%) | 1 (20%) | 4 (40%) | 2 (22.2%) | 0 | 1 (10%) | 0 | 0 | 0 |
| Moderate | 1 (20%) | 0 | 1 (33.3%) | 1 (20%) | 2 (20%) | 2 (22.2%) | 2 (33.3%) | 5 (50%) | 0 | 0 | 0 |
| Severe | 0 | 0 | 1 (33.3%) | 1 (20%) | 0 | 0 | 0 | 0 | 0 | 0 | 0 |

*Supplemental Table 4 continued*

|  | 1μg 3M-052 | | | | 5μg 3M-052 | | | | Placebo | | |
| --- | --- | --- | --- | --- | --- | --- | --- | --- | --- | --- | --- |
| Symptom / Severity | Vacc 1  (N=5) | Vacc 2  (N=4) | Vacc 3  (N=3) | All Vacc (N=5) | Vacc 1 (N=10) | Vacc 2  (N=9) | Vacc 3  (N=6) | All Vacc (N=10) | Vacc 1  (N=2) | Vacc 2  (N=2) | All Vacc (N=2) |
| **Arthralgia** None | 3 (60%) | 4 (100%) | 1 (33.3%) | 2 (40%) | 5 (50%) | 4 (44.4%) | 5 (83.3%) | 3 (30%) | 2 (100%) | 2 (100%) | 2 (100%) |
| Mild | 1 (20%) | 0 | 1 (33.3%) | 1 (20%) | 2 (20%) | 4 (44.4%) | 1 (16.7%) | 4 (40%) | 0 | 0 | 0 |
| Moderate | 1 (20%) | 0 | 0 | 1 (20%) | 3 (30%) | 1 (11.1%) | 0 | 3 (30%) | 0 | 0 | 0 |
| Severe | 0 | 0 | 1 (33.3%) | 1 (20%) | 0 | 0 | 0 | 0 | 0 | 0 | 0 |
| Potentially Life-Threatening | 0 | 0 | 0 | 0 | 0 | 0 | 0 | 0 | 0 | 0 | 0 |
| **Max Systemic Symptoms** None | 0 | 0 | 0 | 0 | 0 | 0 | 0 | 0 | 2 (100%) | 0 | 2 (100%) |
| Mild | 0 | 1 (25%) | 0 | 1 (20%) | 1 (10%) | 1 (11.1%) | 0 | 2 (20%) | 0 | 0 | 0 |
| Moderate | 3 (60%) | 0 | 0 | 3 (60%) | 5 (50%) | 0 | 0 | 5 (50%) | 0 | 0 | 0 |
| Severe | 0 | 0 | 1 (33.3%) | 1 (20%) | 1 (10%) | 0 | 2 (33.3%) | 3 (30%) | 0 | 0 | 0 |
| Potentially Life-Threatening | 0 | 0 | 0 | 0 | 0 | 0 | 0 | 0 | 0 | 0 | 0 |
| **Temperature**  None: < 38C | 5 (100%) | 3 (75%) | 1 (33.3%) | 3 (60%) | 6 (60%) | 7 (77.8%) | 4 (66.7%) | 6 (60%) | 2 (100%) | 2 (100%) | 2 (100%) |
| Grade 1: 38 - < 38.6 C | 0 | 1 (25%) | 2 (66.7%) | 2 (40%) | 3 (30%) | 1 (11.1%) | 0 | 1 (10%) | 0 | 0 | 0 |
| Grade 2: 38.6 - < 39.3 C | 0 | 0 | 0 | 0 | 1 (10%) | 1 (11.1%) | 2 (33.3%) | 3 (30%) | 0 | 0 | 0 |
| Grade 3: 39.3-<40C | 0 | 0 | 0 | 0 | 0 | 0 | 0 | 0 | 0 | 0 | 0 |
| Grade 4: >= 40 C | 0 | 0 | 0 | 0 | 0 | 0 | 0 | 0 | 0 | 0 | 0 |

For a given sign or symptom, each participant's reactogenicity is counted once under the maximum severity for all post-administration assessments at study product administration visits. Symptoms are considered systemic reactogenicity if onset was within 7 days following study product administration.

Maximum systemic symptom is the maximum of the individual variables malaise and/or fatigue, myalgia, headache, nausea, chills, and arthralgia. Reactogenicities are graded according to DAIDS AE Grading Table Version 2.1, July 2017.

1μg 3M-052: BG505 SOSIP.664 gp140 & (1 mcg 3M-052-AF + Alum) month (0,2)

5μg 3M-052: BG505 SOSIP.664 gp140 & (5 mcg 3M-052-AF + Alum) month (0,2)

Placebo for BG505 SOSIP.664 gp140 & adjuvant month (0,2)

**Supplemental Table 5. Summary of antibody and B cell responses among participants with positive BG505 SOSIP.664 binding antibody responses.**

| **3M-052 Dose** | **ID** | **Month 2.5** | | | **Month 6.5** | | |
| --- | --- | --- | --- | --- | --- | --- | --- |
|  |  | **Binding Ab (AUC)** | **Autologous nAb (ID50)** | **BG505 SOSIP gp140 IgG+**  **B cells (%)^#^** | **Binding Ab (AUC)** | **Autologous nAb (ID50)** | **BG505 SOSIP gp140 IgG+**  **B cells (%)^#^** |
| 1 mcg | 137-030 | 3732 | - | 2.802 | 6962 | - | 2.611 |
|  | **137-052** | **14095** | **22.2** | **1.129** | **20626** | **12** | **1.483** |
|  | 137-109 | - | - | 0.764 | 17214 | - | 7.053 |
| 5 mcg | 137-003 | 7878 | - | 2.536 | 17697 | - | 5.733 |
|  | 137-018* | 6410 | - | 3.061 | ND | ND | ND |
|  | 137-116 | 5648 | - | 1.459 | ND | ND | ND |
|  | **137-148** | **6936** | **52.2** | **1.392** | **18029** | **404** | **1.660** |
|  | **137-165** | **12896** | **29.1** | **1.585** | ND | ND | ND |
|  | **137-170** | **14338** | **28** | **6.790** | ND | ND | ND |
|  | **137-185** | **11399** | **-** | **2.608** | **47540** | **8647** | **11.866** |
|  | **137-087** | **2522** | **-** | **2.509** | **23818** | **104** | **5.757** |
|  | 137-214 | 1710 | - | 1.539 | 8821 | - | 4.680 |

- Bold: participants who had positive autologous nAbs at one or more time points
- “-“ designates response below the level of positivity
- ND, not done, designates participants who did not receive a third immunization

*BG505 SOSIP.664 gp41 binding antibodies (AUC) at 2.5 months: 1243

^#^BG505 SOSIP.664 gp140-specific IgG+ B cells of total IgG+ B cells

**Supplemental Table 6. Description of clonotype size, B cell receptor sequence features for B cell-derived monoclonal antibodies from Participant 11.**

| **Clonotype** | **Clonotype size** | **Visit** | **mAb** | **VH Gene** | **VH % mutation** | **HCDR3 length** | **VK/VL Gene** | **VK/VL % Mutation** | **LCDR3 Length** |
| --- | --- | --- | --- | --- | --- | --- | --- | --- | --- |
| 13021_1078 | 32 | 12 | FH31 | IGHV4-30-2*01 | 10.65 | 15 | IGKV2-28*01 or IGKV2D-28*01 | 4.42 | 10 |
| 9503_1102 | 31 | 12 | FH32 | IGHV2-26*01 | 9.62 | 17 | IGKV3-20*01 | 5.32 | 10 |
| 3274_9 | 33 | 12 | FH33 | IGHV3-7*02 or IGHV3-7*04 | 11.46 | 18 | IGKV1-6*01 | 5.73 | 9 |
| 12632_971 | 127 | 12 | FH34 | IGHV1-46*01 or IGHV1-46*03 | 8.42 | 17 | IGKV3-15*01 | 6.45 | 9 |
| 10021_955 | 32 | 12 | FH35 | IGHV2-26*01 | 9.97 | 22 | IGKV3-11*01 | 4.66 | 5 |
| 4516_984 | 40 | 12 | FH36 | IGHV4-31*03 | 10.65 | 18 | IGKV2-28*01 or IGKV2D-28*01 | 4.76 | 9 |
| 12632_971 | 127 | 12 | FH37 | IGHV1-46*01 or IGHV1-46*03 | 11.23 | 17 | IGKV3-15*01 | 6.81 | 9 |
| 7403_434 | 133 | 12 | FH38 | IGHV1-46*01 or IGHV1-46*03 | 13.54 | 18 | IGKV1-17*01 | 4.66 | 9 |
| 9423_502 | 151 | 12 | FH39 | IGHV1-69*02 | 12.50 | 11 | IGKV2-28*01 or IGKV2D-28*01 | 2.38 | 9 |
| 18092_892 | 189 | 12 | FH40 | IGHV3-21*01 | 9.72 | 17 | IGKV1-5*03 | 13.26 | 9 |
| 16368_6911 | 1 | 13 | FH41 | IGHV3-21*06 | 10.07 | 9 | IGKV1-5*03 | 6.45 | 9 |
| 4652_1070 | 169 | 12 | FH42 | IGHV1-46*01 or IGHV1-46*03 | 11.81 | 20 | IGKV1-17*01 | 5.38 | 9 |
| 19155_992 | 46 | 12 | FH43 | IGHV3-23*01 or IGHV3-23D*01 | 11.11 | 14 | IGKV3-11*01 | 4.66 | 10 |
| 15195_25 | 124 | 12 | FH44 | IGHV1-2*02 | 10.42 | 16 | IGKV3-20*01 | 8.87 | 8 |
| 15197_991 | 33 | 12 | FH45 | IGHV1-2*02 | 10.76 | 16 | IGKV3-20*01 | 6.38 | 8 |
